# Latin American Registry of renal involvement in COVID-19 disease. The relevance of assessing proteinuria throughout the clinical course

**DOI:** 10.1101/2021.05.01.21256389

**Authors:** Raúl Lombardi, Alejandro Ferreiro, Daniela Ponce, Rolando Claure-Del Granado, Gustavo Aroca, Yanissa Venegas, Mariana Pereira, Jonathan Chavez-Iñiguez, Nelson Rojas, Ana Villa, Marcos Colombo, Cristina Carlino, Caio Guimarâes, Mauricio Younes-Ibrahim, Lilia Maria Rizo, Gisselle Guzmán, Carlos Varela, Guillermo Rosa-Diez, Diego Janiques, Roger Ayala, Galo Coronel, Eric Roessler, Serena Amor, Washington Osorio, Natalia Rivas, Benedito Pereira, Caroline de Azevedo, Adriana Flores, José Ubillo, Julieta Raño, Luis Yu, Emmanuel A. Burdmann, Luis Rodríguez, Gianny Galagarza-Gutiérrez, Jesús Curitomay-Cruz

**Author notes:** Corresponding author: Raúl Lombardi.

## Abstract

The Latin American Society of Nephrology and Hypertension carried out a cohort prospective, multinational registry of patients with kidney impairment associated to COVID-19 in Latin America through open invitation in order to describe the characteristics of the disease in the region. A population of 870 patients from 12 countries were included. Median age was 63 years (54-74), most of patients were male (68.4%) and had comorbidities (87.2%). Acute kidney injury (AKI) was hospital-acquired in 64.7% and non-oliguric in 59.9%. Multiorgan dysfunction syndrome (MODS) due to COVID-19 and volume depletion were the main causes of AKI (59.2% and 35.7% respectively). Kidney replacement therapy was started in 46.2%. Non-recovery of renal function was observed in 65.3%. 71.5% of patients were admitted to ICU and 72.2% underwent mechanical ventilation. Proteinuria at admission was present in 62.4% of patients and proteinuria during hospital-stay occurred in 37.5%. Those patients with proteinuria at admission had higher burden of comorbidities, higher baseline sCr, higher mortality and MODS was severe. On the other hand, patients with *de novo* proteinuria had lower burden of comorbidities and near normal sCr at admission, but showed adverse course of disease and higher in-mortality. COVID-19 MODS was the main cause of AKI in both groups. All-cause mortality was 57.4%, and it was associated to age, chronic cardiac disease, fluid depletion, COVID-19 MODS, non-recovery of renal function, ICU admission, vasopressors, in-hospital complications and hospital stay. In conclusion, our study contributes to a better knowledge of this condition and highlights the relevance of the detection of proteinuria throughout the clinical course.

## Introduction

Coronavirus disease 2019 (COVID-19), caused by the coronavirus SARS-CoV-2 is an ongoing pandemic that entails high morbidity and mortality rates. Within a month after the first case was reported on February 2020 in Brazil, all countries in Latin America had reported cases of the novel COVID-19 [1]; and by June 2020 Latin America & Caribbean became the worlds’ latest COVID-19 epicenter with the number of deaths in the region exceeding four million, or over 27% of the world’s covid-19 deaths [2]. Sadly, COVID-19 cases are still growing in the region due to constrained health systems, high prevalence of chronic conditions, delayed responses from governments, and widespread poverty and inequality [3]. Growing evidence has demonstrated that kidney involvement, mainly acute kidney injury (AKI) is prevalent among patients with COVID-19, particularly among critically ill patients affecting approximately 20-40% of patients admitted to intensive care units [4, 5]. Similar to the association of AKI with other forms of community-acquired pneumonia [6], AKI is now recognized as a common complication of COVID-19 and as with AKI from other causes, is associated with adverse outcomes [7]. On the other hand, disturbances of the urinary sediment has not been extensively studied in spite its known value. Available information on epidemiology and risk factors for AKI in the region is generally scarce, and this situation has not improved during the COVID-19 pandemic [8]. Knowledge of patient characteristics, risk factors and adverse outcomes, as well as regional peculiarities is key in the fight against this new disease. Taking into account the high incidence of AKI, its high mortality rate and the limited attention given to the associated alterations of the urinary sediment in the region, the AKI Committee of the Latin American Society of Nephrology and Hypertension (SLANH) carried out a prospective cohort study in order to improve the knowledge of this devastating pandemic in Latin America.

## Methods and patients

This is an observational, prospective, longitudinal, multinational cohort study based on a registry carried out between May 1^st^ 2020 and December 31th 2020. An open invitation to participate in the Registry was made through the webpage of the SLANH, the National Societies of Nephrology and by personal email sent to members of *RedIRA* (Latin American AKI-Network) a networked learning tool of SLANH (http://redira.slanh.net/). Participation in the Registry was voluntary, without any incentive or economic benefit for patients or investigators. Data were obtained from the clinical record of patients and were entered online by the participants in a Surveymonkey® platform specifically designed for this purpose (https://es.surveymonkey.com/r/L6PVMGQ). The form has six sections that include: 1) country and city of residence, plus demographic data; 2) comorbidities and condition at admission; 3) laboratory at admission; 4) characteristics and causes of AKI; 5) ICU admission, mechanical ventilation (MV) and in-hospital complications; 6) condition at discharge (S1 Table 1. Form for data colection). Potential bias could exists considering that is a registry of non-consecutive patients. Periodical reminders were sent to participants in order to reduce the rate of non-response and loss of follow-up. We also sent follow-up reminders of incomplete cases throughout the registry with the intention of completing missing cases. Inclusion criteria included adult and children with COVID-19 infection confirmed by RT-PCR of nasopharyngeal swabs and acute kidney injury (AKI) defined by KDIGO 2012 sCr criteria, and/or urinary sediment abnormalities (proteinuria/hematuria). Exclusion criteria: included patients with chronic kidney disease (CKD) stage 5, patients on chronic dialysis or transplanted.

### Definitions

Acute kidney injury was identified according to KDIGO definition when occurred an increase in serum creatinine level ≥ 0.3 mg/dl within 48 hours or by 50% within 7 days. For the detection of proteinuria or hematuria, the semiquantitative dipstick test was used. Renal recovery was considered when sCr returned to baseline or reference value or lower. Non-recovery was established when sCr did not decrease or if the patient remained on dialysis. AKI was considered as community-acquired (CA-AKI) when patient had an elevated SCr within the first 24 hours of admission and hospital-acquired (HA-AKI) when AKI developed during hospital stay.

Condition on admission to the hospital was classified in three categories: mild, if the patient was admitted to a conventional ward without need for oxygen therapy; moderate, if the patient required oxygen therapy; and severe, if the patient was admitted to the intensive care unit (ICU).

Bioethical considerations: the Institutional Review Board of the *Clínica Los Olivos*, Cochabamba, Bolivia (contact Dr. Esdenka Vega) approved the study. The informed consent was considered not mandatory by the reference IRB given the observational characteristic of the study. Protocol and forms are available on the study’s website (https://slanh.net/registro-latinoamericano-ira-covid-19/). Confidentiality of information was appropriately protected by de-identification of data. No personal data of patients was included in the form. Together with the registry, a short survey aimed to know working conditions, difficulties and challenges faced in the course of the pandemic was carried out during the study (S1 Table 2. Results of an open survey of Latin-American nephrologist participants and no participants of the Registry coming from 14 countries. Answers are expressed in percentage).

SLANH provided financial support for the design and maintenance of the platform during the recruitment period, as well as for the publishing fees. The funder had no role in study design, data collection and analysis, decision to publish, or preparation of the manuscript. Competing interests: The authors have declared that no competing interests exist.

### Statistical analysis

Quantitative variables are presented as median and interquartile range (IQR) according to their distribution, and the categorical variables as number and proportions. Kolmogorov-Smirnov test was used to explore the data distribution. For the bivariate comparisons between groups, the Chi square test was used for categorical variables and the *U* Mann-Whitney test or the Kruskal-Wallis test for quantitative variables. Odds ratio and 95% Confidence Interval (CI) were also calculated. Statistical tests were two-sided and significance was considered with a probability of null hypothesis ≤5%. A multiple logistic regression model was performed to predict mortality using the forward conditional elimination method. The probability for entering the model was set at 0.10 and for elimination at 0.20. Variables that reached a significance *<*0.10 in the univariate analysis were included. The statistical package IBM SPSS Statistics Base version 22 NY, USA, was used for data processing and for statistical analysis.

## Results

The overall results of the entire population are presented all together and some subgroups considered of particular interest are described separately.

### 1. Descriptive analysis of the entire population

Forty participants from 52 cities in 12 countries entered 967 patients in the Registry. Ninety-seven patients were excluded due to lack of data on outcomes, remaining 870 patients for the analysis. Median age was 63 (54-74) years; 595 were male (68.4%). Distribution of patients by country is shown in Fig. 1. Patients per country. Of the 870 patients 759 (87.2%) had one or more comorbidities being hypertension, diabetes and obesity prevalent among others (Table 1).

**Fig 1.**
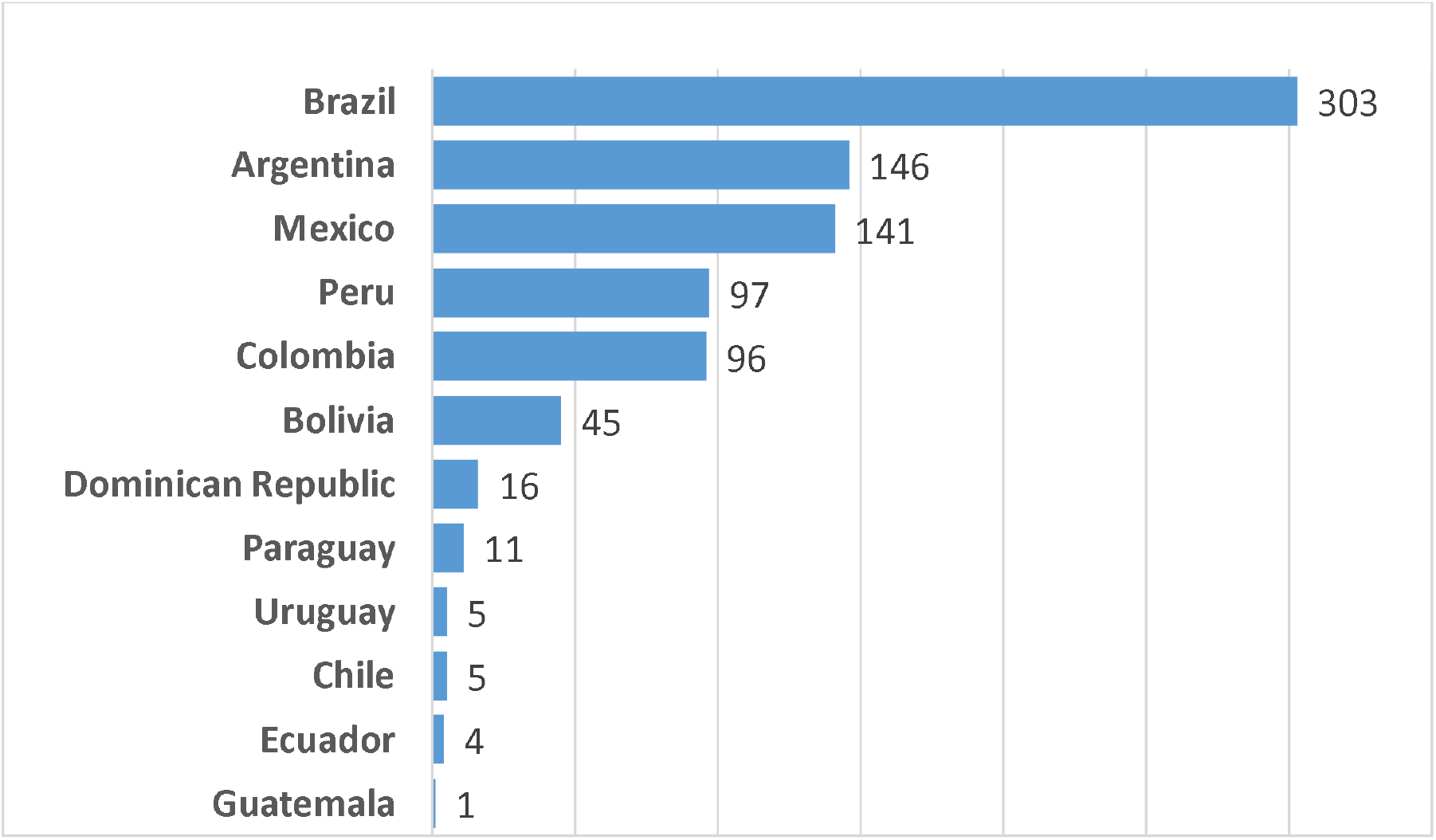
Patients per country

**Table 1.**
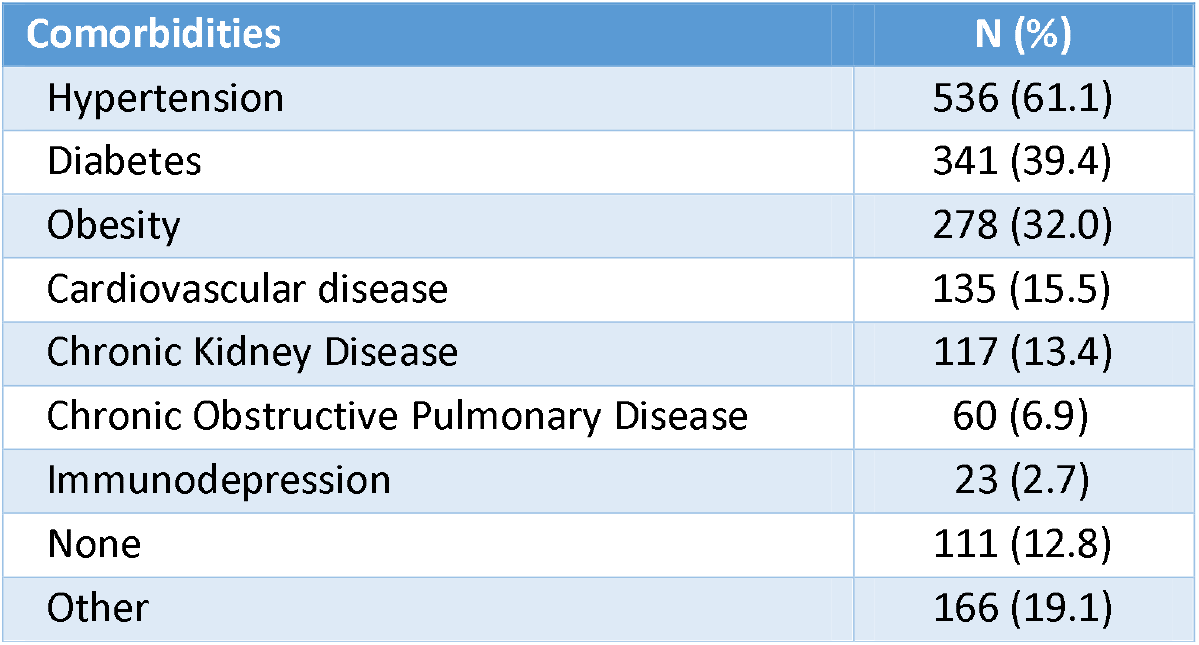
Comorbid conditions

Time between diagnosis of COVID-19 and hospital admission was 2 (0-4) days and condition at hospital admission was mild in 121 (14.0%); moderate in 384 (44.2%) and severe in 363 (41.8%). Table 2 shows laboratory findings at admission and during hospital stay.

**Table 2.** Laboratory data at admission and during hospital stay

| Variable | Median (IQR) |
| --- | --- |
| Scr at admission mg/dL | 1.20 (0.90-1.93) |
| Serum potassium at admission mmol/L | 4.2 (3.9-4.8) |
| WBC at admission mm<sup>3</sup> | 10400 (7800-14137) |
| Platelets x 1000 at admission | 236 (174-302) |
| INR at admission | 1.1 (1.00-1.26) |
| Ferritine at admission (ng/ml) | 1067 (554-1895) |
| pH at admission | 7.39 (7.30-7.43) |
| HCO<sub>3</sub> at admission mmol/L | 22 (19-24) |
| AST at admission (U/L) | 38 (23-63) |
| ALT at admission (U/L) | 32 (20-55) |
| CK at admission (U/L) | 168 (81-421) |
| Scr peak (mg/dL) | 3.60 (1.97-5.20) |
| Minimum PaO<sub>2</sub>/FiO<sub>2</sub> | 134 (112-180) |
| Minimum PEEP (cmH<sub>2</sub>O) | 14 (12-15) |
| Last available Scr (survivors) (mg/dL) | 1.08 (0.81-1.95) |
| Last available Scr (deceased) (mg/dL) | 3.40 (2.11-4.90) |

Of note, about half of patients had a serum creatinine at admission within normal values. In 388 patients a urine dipstick test was performed showing some degree of proteinuria in 242 (62.4%): + 151; ++ 64; +++ 22; ++++ 5. Hematuria was present in 144 out of 390 cases (36.9%). Thirty-nine out of 104 patients (37.5%) developed proteinuria during their hospital stay. AKI was hospital-acquired in 547 (62.9%). Time between diagnosis of COVID-19 and the onset of AKI was 3 (1-7) days. Recorded causes of AKI according to providers’ at each participant center are listed in Table 3 being multi-organ dysfunction (MODS) attributable to SARS-CoV-2 infection the most prevalent.

**Table 3.** Presumed etiology of acute kidney injury, single or in combination

| **Etiology** | **N (%)** |
| --- | --- |
| SARS-CoV-2 MODS* | 515 (59.2) |
| Hypovolemia/dehydration | 311 (35.7) |
| Sepsis MODS | 254 (29.2) |
| Nephrotoxic drugs | 182 (20.9) |
| Rhabdomyolysis | 10 (1.2) |
| Other causes | 110 2.6) |

\*MODS: multiorgan dysfunction syndrome

As expected, in most patients the etiology of AKI was multifactorial or was atributed to more than one exposure. In many patients AKI was linked to more than one etiology. In the majority of cases AKI was non-oliguric (59.9%) and hospital acquired (64.7%). Most patients had KDIGO stage 3 AKI (59.7%) followed by stage 1 (25.8%) and stage 2 (14.5%). Kidney replacement therapy (KRT) was performed in 402 patients (46.2%) and was associated to the level of sCr at admission (OR 1.32, 95% CI 1.00-1.74, p=0.049); admission pH (OR 0.01, 95% CI 0.00-0.44, p=0.016); hospital-acquired AKI (OR 4.26, 95% CI 1.90-9.56, p=0.000); COVID-19 MODS (OR 5.52 95%CI 2.82-10.82, p=0.000); and oliguria (OR 5.97, 95% CI 2.89-12.34, p=0.000). Forty-three patients with indication for KRT didn’t receive it (4.9%) having a higher mortality than the general population (86% vs 62.5%). The reason for not starting KRT was not recorded in each of those cases, but presumably was due to shortage of resources, as can be supported by a contemporary survey answered by 140 Latin American nephrologists 18.3% of them refer having assisted patients who needed KRT but could not receive it (S1 Table 2). The most common type of KRT was intermittent hemodialysis (IHD) (32.4%), followed by prolonged intermittent kidney replacement therapy (PIKRT) (15.2%), continuous kidney replacement therapy (CKRT) (7.2%) and peritoneal dialysis (PD) (1.2%). In the remaining 0.4 % cases other procedures such as hemofiltration and hemoadsorption was performed. Patients were treated for SARS-CoV-2 infection with steroids in 73.9%; oseltamivir in 18.7%; chloroquine-hydroxychloroquine in 12.5%; ivermectine in 6.8%; tocilizumab in 1.5%; remdesivir and linipovir-ritonavir in 0.6%. It should be highlighted that in a large proportion of patients the option “other treatment” was selected in 46.9% with no further clarification. Most of the patients had complications during the hospitalization prevailing sepsis in 50.4% of cases followed by infection without sepsis (8.7%) and deep venous thrombosis (5.4%). Clinical course was severe as expressed by the high number of critically ill patients needing mechanical ventilation (71.5% and 72.2% respectively). As what it was observed with KRT some patients who required ICU and/or mechanical ventilation were not admitted or placed on mechanical ventilation due probably to limited availability of resources in some centers. It should be noted that 6 patients were placed on mechanical ventilation outside the ICU. Renal recovery was observed in 35.2% of patients. All-cause in-hospital mortality was 62.5% (544 out of 870 patients). Of the 544 deceased patients 479 (88.0%) died in ICU. Among survivors, 135 (41.4%) were in ICU and discharged alive from the hospital. Table 4 shows the variables associated to mortality in the univariate analysis and Table 5 the results of the logistic regression model. Variables with more than 30% missing data or inconsistency were excluded from the model (ferritine, platelets count, INR, CK, PaO_2_/FiO_2_).

**Table 4.** Risk factors for in-hospital mortality. Univariable analysis.

| Variable | Death<br>544 | Alive<br>326 | P |
| --- | --- | --- | --- |
| Age yrs | 65 (57-75) | 59 (48-69) | 0.000 |
| Comorbidities n (%) | 94 (17.3) | 41 (12.6) | 0.038 |
| Cardiovascular disease | 199 (36.6) | 79 (24.2) | 0.000 |
| Obesity |  |  |  |
| Condition at admission n (%) | 36 (6.6) | 85 (26.2) | 0.000 |
| Mild | 237 (43.6) | 147 (45.2) |  |
| Moderate | 270 (49.7) | 93 (28.6) |  |
| Severe |  |  |  |
| Scr mg/dL | 1.20 (0.90-1.80) | 1.32 (0.97-2.31) | 0.000 |
| Proteinuria at admission (n 245) | 171 (69.8) | 74 (30.2) | 0.000 |
| White blood cell count mm3 | 11200 (8225-15795) | 9559 (6975-12710) | 0.000 |
| Ferritine ng/ml | 1285 (689-2000) | 840 (366-1307) | 0.000 |
| CK U/L | 228 (99-669) | 123 (60-250) | 0.000 |
| Hospital-acquired AKI n (%) | 399 (74.0) | 148 (48.2) | 0.000 |
| Cause of AKI n (%) |  |  |  |
| Hypovolemia | 156 (28.7) | 155 (47.59) | 0.000 |
| MODS* SARS-CoV2 | 376 (69.1) | 139 (42.6) | 0.000 |
| MODS sepsis | 207 (38.1) | 47 (14.4) | 0.000 |
| Nephrotoxic drugs | 99 (18.2) | 83 (25.5) | 0.007 |
| Oliguric AKI | 259 (48.3) | 77 (25.0) | 0.000 |
| Scr peak mg/dL | 4.10 (2.80-5.47) | 2.15 (1.49-4.40) | 0.000 |
| Kidney replacement therapy | 315 (58.0) | 87 (27.0) | 0.000 |
| Non-recovery of renal function | 462 (88.5) | 71 (23.7) | 0.000 |
| Duration of kidney replacement therapy (days) | 6 (3-11) | 10 (5-27) | 0.000 |
| Admission to ICU | 484 (89.3) | 138 (42.3) | 0.000 |
| Mechanical ventilation | 492 (90.6) | 136 (42.1) | 0.000 |
| Minimum PaO<sub>2</sub>/FiO<sub>2</sub> | 123 (108-157) | 164 (123-237) | 0.000 |
| New onset of proteinuria | 106 (32.8) | 57 (24.1) | 0.015 |
| New onset of hematuria | 101 (30.9) | 32 (13.7) | 0.000 |
| Last available Scr mg/dL | 3.40 (2.10-4.90) | 1.07 (0.82-1.90) | 0.000 |
| Complications (%) |  |  | 0.000 |
| Sepsis | 360 (67.2) | 79 (25.1) | 0.000 |
| Infection | 22 (4.3) | 54 (17.4) | 0.000 |
| No complications | 51 (9.9) | 110 (34.9) |  |
| Hospital stay (days) | 13 (8-21) | 14 (8-28) | 0.046 |

\*MODS: multiorgan dysfunction syndrome

**Table 5.**
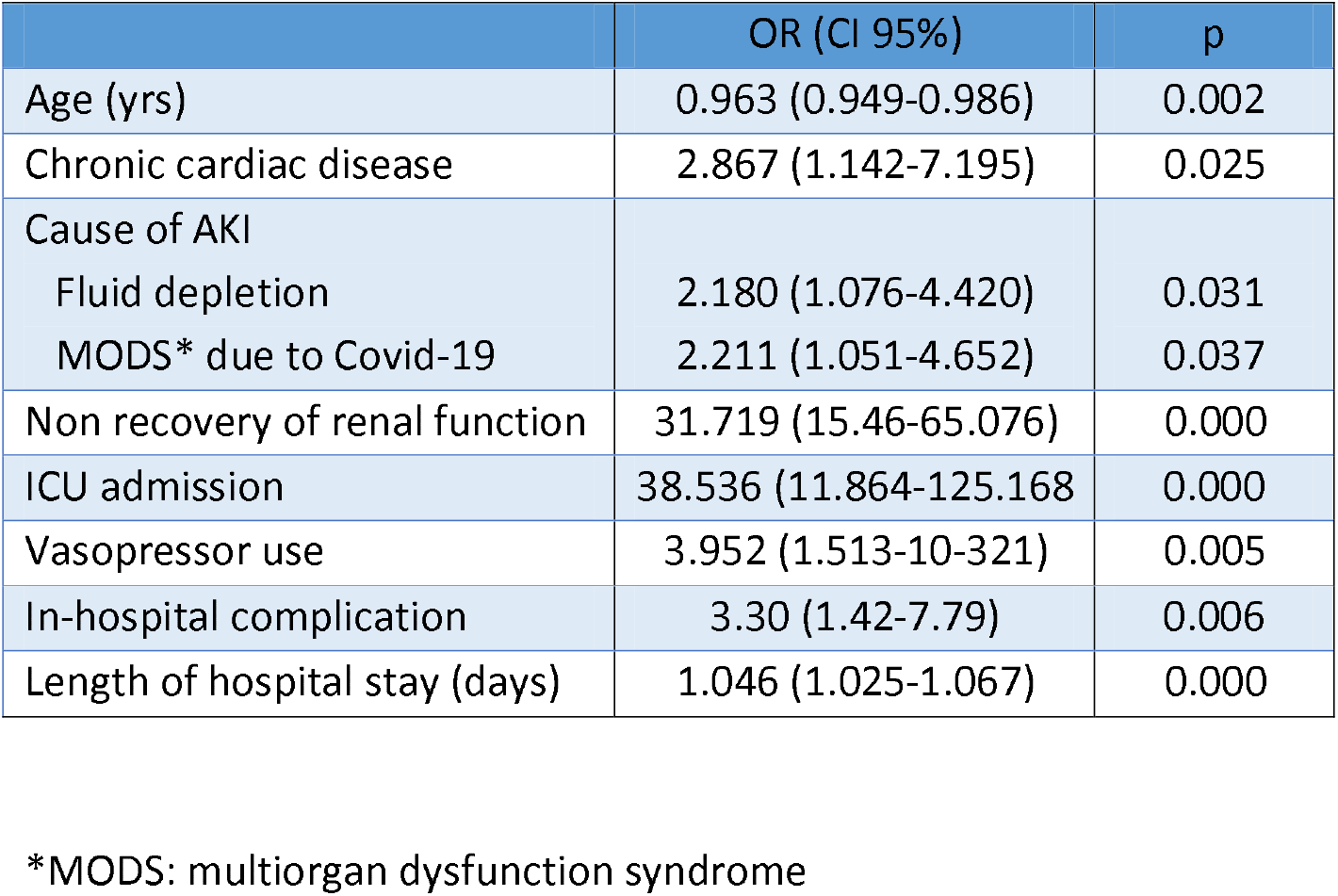
Risk factors independently associated to in-hospital mortality. Multivariable logistic regression analysis.

#### Subgroup I. Patients with proteinuria on admission

Characteristics of subgroup I are shown in Table 6.

**Table 6.** Clinical characteristics of patients with proteinuria at admission. Subgroup I.

| Variable | Proteinuria<br>242 | No proteinuria<br>146 | p |
| --- | --- | --- | --- |
| Comorbidities n (%) |  |  |  |
| Hypertension | 165 (67.3) | 85 (57.4) | 0.039 |
| Chronic kidney disease | 54 (22.0) | 7 (4.7) | 0.000 |
| No comorbidities | 23 (9.4) | 24 (16.2) | 0.032 |
| Cause of acute kidney injury |  |  |  |
| SARS-CoV-2 MODS | 162 (66.1) | 83 (56.1) | 0.030 |
| Scr at admission mg/dl | 1.39 (0.90-2.64) | 0.99 (0.80-1.30) | 0.000 |
| Serum potassium at admission | 4.35 (4.00-5.02) | 4.10 (3.80-4.40) | 0.000 |
| Scr peak mg/dL | 4.10 (2.70-5.55) | 3.00 (1.50-4.90) | 0.000 |
| Kidney replacement therapy | 136 (56.0) | 60 (40.8) | 0.006 |
| Renal function recovery | 59 (25.5) | 61 (47.7) | 0.000 |
| ICU admission | 195 (79.9) | 100 (67.6) | 0.02 |
| Mechanical ventilation | 198 (81.1) | 101 (68.2) | 0.013 |
| Vasopressors | 158(67.2) | 74 (53.2) | 0.005 |
| In-hospital complication<br>Infection | 23 (9.4) | 33 (22.7) | 0.001 |
| Last available Scr mg/dL | 3.00 (1.60-4.50) | 1.56 (0.90-3.14) | 0.000 |
| Mortality | 171 (69.8) | 74 (50.0) | 0.000 |

In summary, this subgroup of patients had higher burden of comorbidities, and elevated sCr and potassium at admission. AKI was linked to COVID-19 MODS and showed worse outcomes (severity of the disease and mortality). Within the Subgroup I, 34 out of 393 patients did not developed AKI. As expected those cases had milder forms of COVID-19 at admission and had a less severe clinical course during hospitalization (need of ICU, MV, vasopressors, and mortality).

### 2. Subgroup II. Patients with normal urinary sediment on admission and *de novo* proteinuria during hospital stay

Of 104 patients in whom urinary sediment was assessed during their hospitalization, 39 (37.5%) of them developed *de novo* proteinuria. Clinical characteristics are shown in Table 7.

**Table 7.**
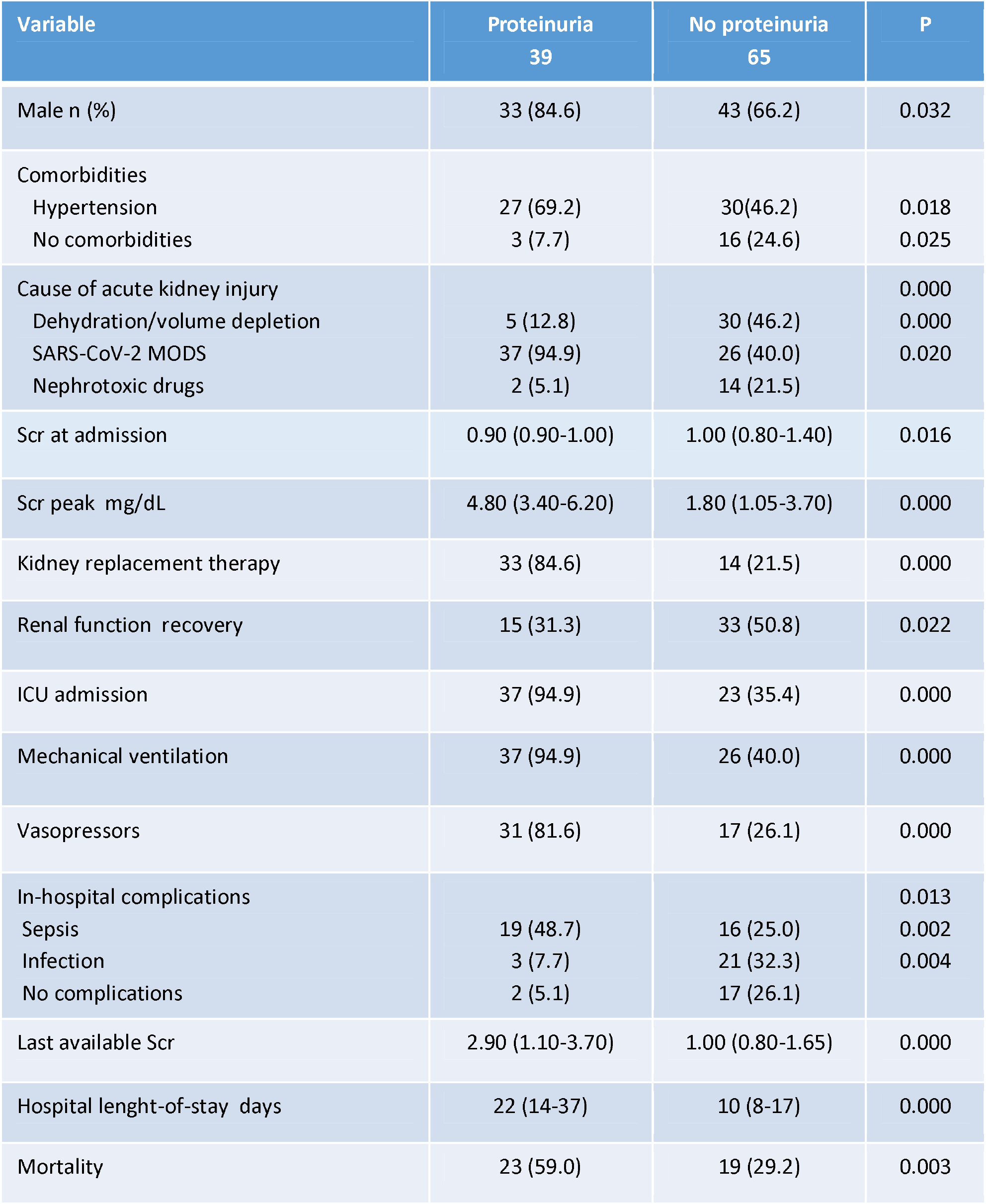
Characteristics of patients who developed *de novo* proteinuria. Subgroup II.

In brief, prevalence was higher in males, the sCr at admission was normal and the predominant cause of AKI was MODS associated to COVID-19. The clinical course was characterized by higher AKI stages, KRT requirement, higher incidence of organ dysfunction and complications, less recovery of renal function and higher mortality.

### 3. Critically ill patients

A large proportion of patients needed to be admitted to the ICU (622, 71.5%) and 628 (72.2%) were placed on mechanical ventilation. Table 8 shows the different characteristics between critically ill and no critically ill patients.

**Table 8.** Clinical characteristics by severity of clinical condition. Subgroup critically ill patients.

| Variable | ICU<br>622 | No ICU<br>221 | P |
| --- | --- | --- | --- |
| Obesity n (%) | 227 (35.5) | 40 (18.1) | 0.000 |
| Proteinuria at admission | 195/295 (66.1) | 46/91 (50.5) | 0.023 |
| Hospital-acquired acute kidney injury | 445/620 (71.8) | 90/201 (44.8) | 0.000 |
| Cause of acute kidney injury |  |  |  |
| SARS-CoV-2 MODS* | 432 (69.5) | 66 (29.9) | 0.000 |
| Sepsis MODS | 229 (36.8) | 19 (8.6) | 0.000 |
| Nephrotoxic drugs | 113 (18.2) | 65 (29.4) | 0.002 |
| Days COVID-hospital admission | 2 (1-5) | 1 (1-3) | 0.000 |
| Scr at admission | 1.10 (0.89-1.61) | 1.50 (1.10-2.67) | 0.000 |
| White blood cells count mm<sup>3</sup> | 10700 (7800-14700) | 9600 (7295-13397) | 0.028 |
| Ferritine ng/mL | 1291 (750-2000) | 647 (349-1190) | 0.000 |
| pH | 7.39 (7.30-7.42) | 7.41 (7.30-7.45) | 0.004 |
| HCO<sub>3</sub> mmol/L | 22 (19-24) | 21 (18-24) | 0.034 |
| Days COVID-AKI onset | 4 (2-9) | 1 (1-3) | 0.000 |
| Last available Scr mg/dL | 3.00 (1.43-4.40) | 1.20 (0.90-2.20) | 0.006 |
| Mortality | 314 (50.6) | 35 (15.8) | 0.000 |

Obesity, proteinuria, hospital acquired AKI and MODS related AKI, delay in hospital admission, higher values in white blood cells and ferritine and, as expected, higher mortality was the profile of this subset of patients.

## Discussion

In early December 2019, an outbreak of an acute respiratory infection caused by SARS-CoV-2 virus occurred in Wuhan City, China, which rapidly spread to other regions of China and later globally constituting a pandemic as was declared by WHO on March 11, 2020. The respiratory system is the primary target of the virus, but it was increasingly recognized that it could affect other organs, including kidneys leading to proteinuria, haematuria and AKI [9,10, 11]. On the other hand, kidney impairment and mortality increases with disease severity [5,12,13]

The real incidence of AKI in COVID-19 is cause of debate. Early studies from China showed low AKI incidence (0.5%) in a series of 1,099 hospitalized patients. [14]. Conversely, a large series of 5,449 patients admitted to 13 academic and community hospitals in New York, identified 1,993 patients with AKI (36.6%) [5], an incidence close to the 33.9% reported in 35,302 patients from the largest hospital discharge database in USA (Premier Healthcare Database) [15]

Currently, America is the epicenter of COVID-19 pandemic being the impact devastating on the Latin America subcontinent [16,17,18]. The rapid spread of the infection, the work overload of health teams, the uneven level of health systems in the region, and the differences in strategies in the fight against the pandemic between countries, partly explains the lack of data in a region, that otherwise has scarce general epidemiological information on AKI [19].

In order to improve gaps in the knowledge of kidney disorders associated with COVID-19 in the region the AKI Committee of the Latin American Society of Nephrology and Hypertension conducted a Registry that gathered data of 870 patients from 57 cities distributed in 12 countries in which the elderly male prevailed. Our population showed a high burden of comorbidities, since 87.2% had at least one associated disease, mainly hypertension, diabetes and obesity. Moreover, severe clinical condition at hospital admission was also predominant (86%). Finally, time elapsed between diagnosis of COVID-19 and hospitalization was short, what can be read as a marker of severity of the disease. It is not surprising therefore, that in-hospital mortality was high (62.5%), greater than reported in other studies [5,12]. In contrast with the severity of SASR-CoV-2 infection at admission, renal and metabolic condition appeared to be less affected. In most of the cases, sCr level was near normal, as well as serum potassium and acid-base status. However, evidence of ongoing kidney impairment at admission was demonstrated since in 62.4% of the patients in whom a urine dipstick test was performed had proteinuria. With regard to AKI profile, the onset occurred shortly after COVID-19 diagnosis and was linked in the vast majority of cases to COVID-19 MODS and hypovolemia. As previously reported, the onset of AKI was mainly during hospital stay prevailing a non-oliguric pattern [5] and severe forms. An additional burden of seriousness was due to in-hospital complications, mostly sepsis (50.4%).

In previous studies variable degrees of proteinuria were reported among COVID-19 patients, ranging between 33% and 74% with a higher incidence among AKI stages 2-3 (11) and it was invariably associated to worse outcome [5,9,]. Histopathological studies showed a diversity of presentations like collapsing glomerulopathy, thrombotic microangiopathy, minimal change disease, immune-mediated glomerulopathy and acute tubular necrosis (20,21). We assessed the presence of proteinuria at admission and through patients’ hospital stay (*de novo* proteinuria). By doing that, we identified two patterns of proteinuria having interesting different characteristics between them. Group I patients’ (proteinuria at hospital admission) had more comorbidities, were admitted with impaired renal function, had a worse course of their illness leading to a significant rise in mortality rate, as can be seen in Table 5. On the other hand, Group II patients (*de novo* proteinuria) had normal kidney function at hospital admission, which would anticipate a better outcome. However, in most of the cases their clinical condition worsened requiring multi-organ support, recovered renal function less frequently, had prolonged hospital stay and a higher mortality rate (Table 6). To our knowledge, the latter pattern of proteinuria that portends poor results has not been reported previously being one of the most relevant findings of our study. In both groups, SARS-CoV-2 MODS was recognized by far as the main cause of AKI that strongly suggests glomerular and tubular damage due to severe inflammation, together with associated hemodynamic disturbances. These results lead us to strongly recommend including a urine dipstick test in the routine evaluation of COVID-19 patients for a timely detection of kidney injury.

As previously reported, AKI is associated to higher risk of death in COVID-19 patients [5,15,20]. Our study found higher than usual mortality rate, probably due to a selection bias since our Registry is an open repository of patients provided by selected physicians (nephrologists). We found as many as 24 variables associated with mortality in the univariate analysis. Unfortunately, some of them could not be included in a regression model because of inconsistence of data or high number of patients with missing data. With these limitations, we identified nine variables that were independently associated to in-hospital mortality. Older age (OR 0.96, 95% IC 0.94-0.99); heart disease (OR 2.87, 95% IC 1.14-7.19); fluid depletion (OR 2.18, 95% IC 1.08-4.42) and COVID-19 MODS (OR 2.21, 95% IC 1.05-4.65) as cause of AKI; non-recovery of renal function (OR 31.72, 95% IC 15.46-65.08); critically ill condition (OR 38.54, 95% IC 11.86-125.17); use of vasopressor (OR 3.95, 95% IC 1.51-10.32); some in-hospital complications (OR 3.30, 95% IC 1.42-7.79) and hospital length-of-stay (OR 1.05, 95% IC 1.02-1.07). Of note, sCr requested at three time points (admission, peak, discharge) did not enter in the model.

Our study has some limitations. First, its design as a repository of patients promote the individual participation in addition to that of institutions, as desired. In return, entail some heterogeneity in the quality of information resulting in some variables having a high proportion of missing data that forced us to exclude them from the analysis. Consequently, potential unmeasured confounder may not have been identified. Second, the limited ability to access the universe of millions patients did not allow us to establish incidence, which is a very relevant information, particularly considering that is a controversial and open point. Third, data of patients was provided by the participants and not as a result of the review of clinical charts by the research team. We also acknowledge some strengths. First, to date this is the largest multicenter study involving a large number of countries and cities from Latin America. Second, data requested were organized in six forms with 47 questions with the aim to fully explore the more relevant problems related to SARS-CoV-2 infection and kidney disease. With this approach, two different patterns of patients with proteinuria having relevant prognostic value were clearly identified.

In conclusion, this is the first multinational Registry of patients with renal impairment associated to SARS-CoV-2 infection in Latin America. The data of 870 patients from 14 countries generally shows similar disease profile to previously reported series regarding to demographic, underlying condition, AKI features, impact on distant organs and mortality. The study highlights the value of assessing proteinuria throughout the clinical course for a timely identification of renal impairment and potential adverse outcomes. Our results cannot be generalized, as it is a repository of patients that partially represents the universe of this condition. However, this study demonstrates the feasibility of carrying out similar studies in order to expand the knowledge of this new disease that remains to be fully characterized.

## Data Availability

Data cannot be shared publicly because of ethical restrictions. In spite cases are
appropriately deidentified data are available only on request. Contact via
Powered

https://slanh.net/registro-latinoamericano-ira-covid-19/

## Acknowledgments

We would like to thank all the clinical staff and the hospitals that were part of this 57 collaboration in 12 countries, for supporting this project.

## Authors contribution

Conceptualization and design: RL, AF, RCD, MYI, LY, EAB

Data curation: RL

Acquisition of data, analysis and interpretation: all authors

Drafting the manuscript: RL, AF, DP, RCD

