## Supplementary material for "Latin American Registry of renal involvement in COVID-19 disease. The relevance of assessing proteinuria throughout the clinical course": Table 1 and 2

**Supplementary Table S1. Form for data collection**

*1. Demographic data and location*

a. Age

b. Sex

c. City / town of residence

d. Country

*2. Comorbidities and condition at admission*

a. Comorbidities (hypertension, diabetes, obesity, chronic heart failure, chronic kidney disease, chronic obstructive pulmonary disease, immunodepression)

b. Days between COVID-19 diagnosis and admission to hospital

c. Hospital admission date

d. Condition at admission (mild, moderate severe)

*3. Laboratory at admission*

i. Serum creatinine

ii. Potasemia

iii. Proteinura at admission and *de novo* during hospital stay

iv. Hematuria

v. White blood cells count

vi. Lymphocytes count

vii. Platelets count

viii. INR

ix. D-dimers

x. Ferritin

xi. pH

xii. HCO3

xiii. ALT

xiv. AST

xv. CK

xvi. PaO2

*4. Characteristics and cause of AKI*

a. Community acquired/Hospital acquired

b. Days between COVID-19 and diagnosis of AKI

c. Etiology of AKI

i. Fluid depletion/shock

ii. Multiorgan dysfunction syndrome due to sepsis or pro-anti-inflammatory cascade

iii. Rhabdomyolysis

iv. Nephrotoxic drugs / contrast

v. Others	id. Diuresis (oliguric / non-oliguric)

d. Serum creatinine peak

e. Kidney replacement therapy (yes/no)

i. Conventional HD

ii. SLED

iii. Continuous KRT

iv. HDF online

v. Hemoadsorption

vi. DP

f. The patient had indication of KRT but did not receive it (yes/no)

g. Recovery of renal function (yes / no)

h. Duration of AKI (if recovered)

*5. Process of care*

a. ICU admission (yes / no)

b. Days in ICU

c. The patient had ICU indication but was not admitted

d. Invasive mechanical ventilation

e. Worst Pa / FiO2

f. Worst PEEP

g. The patient had indication of mechanical ventilation but was not performed

*6. Condition at hospital discharge*

a. Dead

b. Alive discharge from hospital

c. Transferred to another hospital

d. Scr at discharge

e. Hospital length-of stay

**Supplementary Table S2. Results of an open survey of Latin-American nephrologist participants and no participants of the Registry coming from 14 countries. Answers are expressed in percentage**

| Gender  Male  Female | 46.4  53.6 |
| --- | --- |
| Position of responders  Head of department  Nephrologist (ward)  Intensivists  Other | 19.3  68.5  2.9  9.3 |
| Did you modify your activity during the pandemic because being a risk group for COVID-19?  Yes  No | 35  65 |
| Did you modify your activity during the pandemic not being risk group for COVID-19 by your own?  Yes  No | 5  95 |
| Did you was regularly provided by PPE in your hospital?  Yes  No  Occasionally | 60.9  34.1  5.0 |
| Did you always had the necessary supplies to perform KRT in patients with any cause AKI?  Yes  No  Occasionally | 53.3  16.0  30.7 |
| Did you assist COVID-19 patients needing KRT but who did not receive it?  Very frequently  Frequently  Occasionally  No | 9.5  8.8  27.0  54.7 |
| Did you assist COVID-19 patients needing intensive care that were not admitted in ICU?  Yes  No | 27.3  72.7 |
| Did you assist COVID-19 patients needing mechanical ventilation but who did not receive it?  Yes  No | 21.6  78.4 |
| In your hospital, regular dialysis schedule was modified in order to increase the renal replacement capacity for patients with any cause AKI?  Yes  No | 38.1  61.9 |
| In your hospital, was PD incorporated or increased for the treatment of patients with any cause? AKI  Yes  No | 10.1  89.9 |
| Has a health care worker of your hospital been infected with COVID-19 because of their practice?  Yes  No | 77.9  22.1 |
| Has a health care worker of your hospital died of COVID-19 infection?  Yes  No | 39.3  60.7 |
